# Seroprevalence of SARS-CoV-2 antibodies in social housing areas in Denmark

**DOI:** 10.1101/2021.05.07.21256725

**Authors:** Kamille Fogh, Alexandra RR Eriksen, Rasmus B Hasselbalch, Emilie Sofie Kristensen, Henning Bundgaard, Susanne D Nielsen, Charlotte S Jørgensen, Bibi FSS Scharff, Christian Erikstrup, Susanne G Sækmose, Dorte K Holm, Bitten Aagaard, Jakob Norsk, Pernille Brok Nielsen, Jonas H Kristensen, Lars Østergaard, Svend Ellermann-Eriksen, Berit Andersen, Henrik Nielsen, Isik S Johansen, Lothar Wiese, Lone Simonsen, Thea K. Fischer, Fredrik Folke, Freddy Lippert, Sisse R Ostrowski, Steen Ethelberg, Anders Koch, Anne-Marie Vangsted, Tyra Grove Krause, Anders Fomsgaard, Claus Nielsen, Henrik Ullum, Robert Skov, Kasper Iversen

## Abstract

**Background:** COVID-19 is suggested to be more prevalent among ethnic minorities and individuals with low socioeconomic status. We aimed to investigate the prevalence of SARS-CoV-2 antibodies during the COVID-19 pandemic among citizens 15 years or older in Denmark living in social housing (SH) areas.

**Methods:** As part of “Testing Denmark”, a nationwide sero-epidemiological surveillance survey, we conducted a study between January 8^th^ and January 31^st^, 2021 with recruitment in 13 selected SH areas in Denmark. Participants were offered a point-of-care rapid SARS-CoV-2 IgM and IgG antibody test and a questionnaire concerning previous testing (viral throat- and nasopharyngeal swab or antibody test), test results for COVID-19, demographics, household characteristics, employment, risk factors for SARS-CoV-2 infection and history of symptoms associated with COVID-19. Data on seroprevalence from Danish blood donors in same period using a total Ig ELISA assay were used as a proxy for the general Danish population.

**Findings:** Of the 13,279 included participants, 2,296 (17.3%) were seropositive (mean age 46.6 (SD 16.4) years, 54.2% female), which was 3 times higher than in the general Danish population (mean age 41.7 (SD 14.1) years, 48.5% female) in the same period (5.8%, risk ratios (RR) 2.96, 95% CI 2.78-3.16, p>0.001). Seropositivity was higher among males than females (RR 1.1, 95% CI 1.05-1.22%, p=0.001) and increased with age, with an OR seropositivity of 1.03 for each 10-year increase in age (95% CI 1.00-1.06, p=0.031). Close contact with COVID-19-infected individuals was associated with a higher risk of infection, especially among members of the same households (OR 5.0, 95% CI 4.1-6.2 p<0,001). Adjusted for age, gender and region living at least 4 people in a household significantly increased the OR of seropositivity (OR 1.3, 95% CI 1.1-1.6, p=0.02) as did living in a multi-generational household (OR 1.3 per generation, 95% CI 1.1-1.5, p=0.007). Only 1.6% of participants reported not following any of the national COVID-19 recommendations. Anosmia (RR 3.2 95% CI 2.8-3.7, p<0.001) and ageusia (RR 3.3, 95% CI 2.9-3.8, p<0.001) were strongest associated with seropositivity.

**Interpretation:** Danish citizens living in SH areas of low socioeconomic status had a three times higher SARS-CoV-2 seroprevalence compared to the general Danish population. The seroprevalence was significantly higher in males and increased with age. Living in multiple generations or more than four persons in a household was an independent risk factor for being seropositive. Results of this study can be used for future consideration of the need for preventive measures in the populations living in SH areas.

## Introduction

The first confirmed case of SARS-CoV-2 infection in Denmark was reported on February 27, 2020 and by May 4^th^, 2021 there have been more than 254,482 confirmed cases of SARS-CoV-2 infection and more than 2491 COVID-19 related deaths in Denmark (1).

So far, the outbreak of the epidemic has had a heterogeneous regional patterns with geographical accumulations and varying incidence by gender, age, social class and employment (2). Although there is equal and free of charge access to health care for everybody in Denmark including testing for COVID-19 (viral throat- and nasopharyngeal swab), citizens’ behavior may vary in different social segments. National and regional seroprevalence data offer valuable information to tailor public health policies towards the COVID-19 epidemic.

According to the Danish authorities, 15 residential areas are currently defined as social housing (SH) areas, characterized by low employment rates, low income, low education level, high crime rate and/or increased proportion of immigrants (3). Some reports suggest that ethnic minorities in a number of countries are over-represented among those infected with COVID-19, just as socioeconomic inequality is described among SARS-CoV-2 infected persons (4-6). A Danish report from October 2020 showed similar patterns, where people of non-Western background accounted for 25.7% of cases with SARS-CoV-2 infection, despite representing only 8.9% of the population (7, 8).

Vulnerable and marginalized populations, certain ethnic minorities and persons of low socioeconomic status may have difficulties receiving and following health recommendations (9). Which could lead to reduced use of protective equipment and difficulties in navigating the health care system with impaired contact, due to cultural and linguistic barriers, with the risk of being underdiagnosed. For cultural and economic reasons, individuals in SH areas may live in crowded multi-generational households with children, parents and grandparents, which has been hypothesized to increase transmission of SARS-CoV-2 (4, 10). This may not only affect their households but also people in their environment. Estimating the contributions of individual and sociocultural factors that may lead to COVID-19 outbreaks is important, and systematic screening for antibodies against SARS-CoV-2 is an important tool in the surveillance of the current pandemic.

The Danish prevalence of SARS-CoV-2 seropositivity is reported for blood donors (11), medical students (12) hospital staff (13) and in a random sample of Danish citizens (14), but not in a subpopulation that may be at increased risk of SARS-CoV-2 infection due to low socioeconomic status.

In this study we determined the prevalence of SARS-CoV-2 antibodies among Danish citizens in SH areas at national and regional levels, by the use of Point-of care rapid test (POCT) for antibodies against SARS-CoV-2 and explored possible risk factors of seropositivity.

## Methods

### Study design and participation

The sero-epidemiological survey of SARS-CoV-2 infection in Denmark “Testing Denmark” is a nationwide surveillance study to investigate seropositivity for SARS-CoV-2 in the Danish population throughout the country, launched in September 2020.

The prevalence of SARS-CoV-2 antibodies among Danish citizens in SH areas was assessed by use of POCT during the period January 8^th^ and January 31^st^, 2021 as part of “Testing Denmark”.

Recruitment sites were chosen in collaboration with non-governmental organization with an ethnic minority background, who do voluntary efforts in their local community area. We recruited participants from 13 different SH areas in Denmark (see appendix, Figure 1). Persons more than 15 years of age were invited to participate.

**Figure 1:**
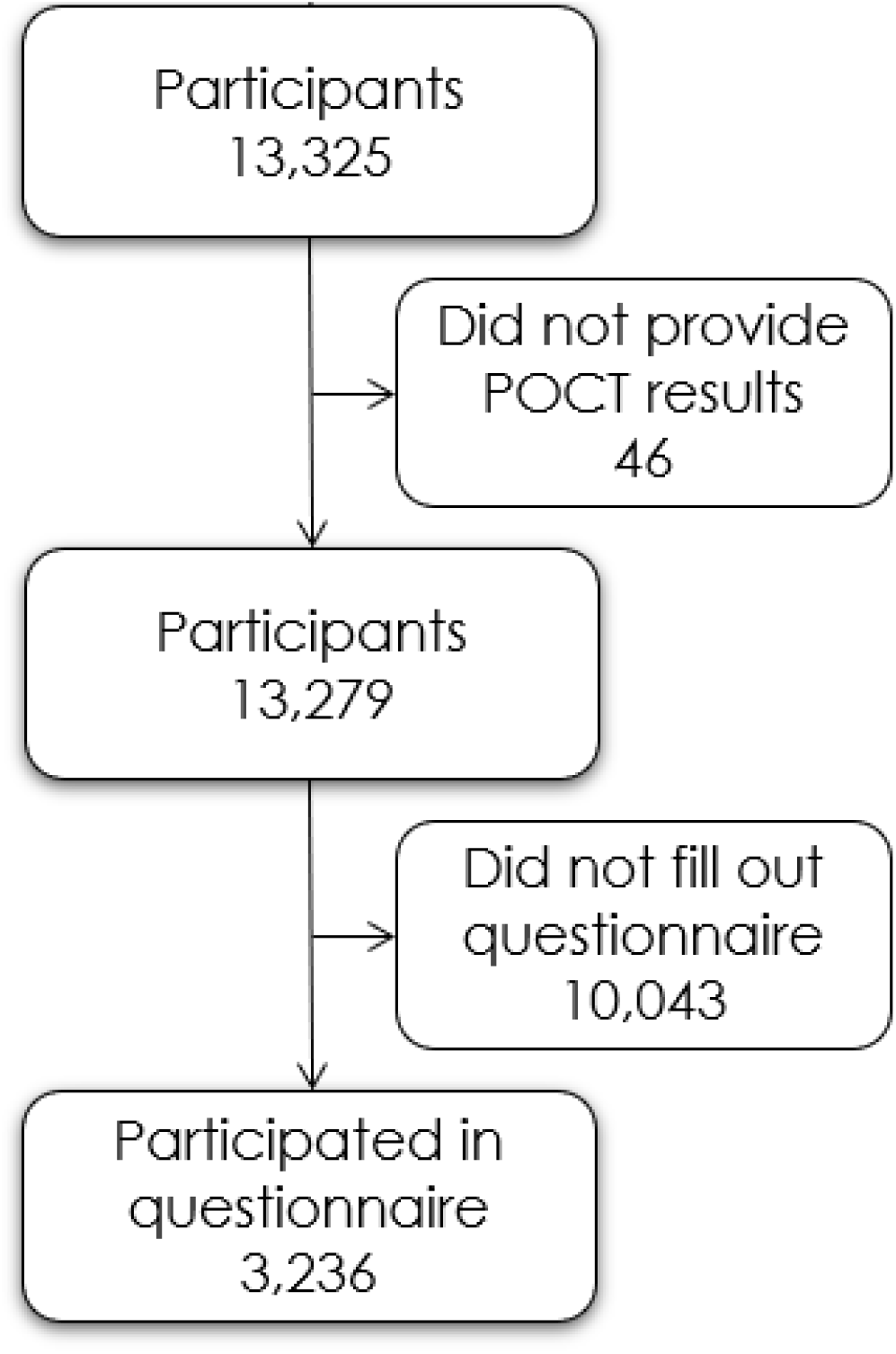
CONSORT diagram

At all recruitment sites written information about the project were available in seven different languages; Danish, English, Arabic, Turkish, Farsi, Somali and Urdu.

### Questionnaire

Participants were asked to fill in a short questionnaire provided at the recruitment site, available in the seven different languages, due to risk factors, COVID-19 related symptoms, household, employment and behavior according to general recommendations from the Danish authorities (see questionnaire in appendix).

Answers to the questionnaire and results of the POCT were managed in the Research Electronic Data Capture (REDCap), a secure web-based, electronic data capture tool, hosted at the Capital Region’s server (15, 16). All personal data was kept in accordance with the general data protection regulation and data protection law stated by the Danish Data Protection Agency.

### Detection of SARS CoV-2 antibodies

The OnSite COVID-19 IgG/IgM Rapid Test (CTK Biotech inc., Poway, California, United States of America) is a single use lateral flow chromatographic immunoassay for qualitative detection and differentiation of IgG and IgM antibodies to SARS-CoV-2 in whole blood, which yields results in 15 minutes. This test was used by the participant with assistance from the project personnel, according to the manufacturer’s recommendations.

The manufacturer reported the test sensitivity and specificity of 96.9% (95% CI: 96.7% - 98.5%) and 99.4% (95% CI: 97.8% - 99.8%) respectively (17). A comparative study (cases=30 individuals, controls=10 individuals) showed a slightly lower sensitivity of 90.0% and a specificity of 100% (18).

Fingerprick blood and detection buffer were added to the test cassette and test results were available after 15 minutes by trained project personnel. When no control line appeared or if the reading chamber was discolored by blood the test was inconclusive. Inconclusive test results were treated as negative. Weak signals for IgM and/or IgG, were considered positive. Participants were categorized as seropositive if they had developed IgG and/or IgM anti-SARS-CoV-2 antibodies.

### SARS-CoV-2 antibody levels in the general population

Since October 2020, all Danish blood donations are routinely screened for SARS-CoV-2 antibodies using the Wantai SARS-CoV-2 Ab ELISA (Beijing Wantai Biological Pharmacy Enterprise, Beijing, China). In this study we used anonymized data from January 2021, matched by period. This group was used as a proxy for the general population.

### Primary outcome

The primary outcome was the proportion of the study population with a positive antibody test for SARS-CoV-2 stratified by place of testing compared to the general population.

### Approvals and registrations

This study was performed as a national cross-sectional surveillance study under the authority task of Statens Serum Institut (SSI, Copenhagen, Denmark; which performs the epidemiological surveillance of infectious diseases for the Danish government) and according to Danish law does not require any formal approval from an ethics committee. This was in accordance with the regional ethics committee from the Capital region in Denmark (20057075). The study was performed in agreement with the Helsinki II declaration and registered with the Danish Data protection Authorities (P-2020-901). Participation was voluntary. All personal data obtained in REDCap was kept in accordance with the general data protection regulation and data protection law stated by the Danish Data Protection Agency.

### Statistical analysis

Baseline characteristics and exposures are presented as n (%) for factors and mean (standard deviation (SD) or median (interquartile range (IQR)) for numeric variables as appropriate. Household size was presented both as the total number of persons (with a maximum of >5 according to the questionnaire) and as the number of generations in the household. The three generations were defined as individuals per household; <19, 19-64 and >65 years of age.

Unadjusted risk was presented as risk ratios (RR) with 95% confidence intervals (95% CI). To account for the possible clustering effect of participants, we chose to use logistic regression adjusting for test location (SH area) to determine the correlation between putative risk factors and seropositivity. Results of these regressions analyses were reported as odds ratios (OR) of risk factors and presented with 95% confidence intervals (95% CI). A p-value of <0.05 was considered significant. Calculation were done using R version 6.3.1 (19).

## Results

### Characteristics

Between January 8^th^ and January 31^st^, 2021, we included a total of 13,279 participants in SH areas. The mean age of the cohort was 46.6 (SD 16.4) years and 54.2% were female. Baseline characteristics of the cohort is shown in table 1. A total of 3,236 (24.4%) completed the accompanying questionnaire, primarily in Danish (94.4%). A flowchart of participant inclusion is depicted in figure 1.

**Table 1:** Baseline characteristics of the study cohort of people in SH areas stratified by seropositivity

| Full cohort | Seronegative | Seropositive | p |
| --- | --- | --- | --- |
| n | 10,983 | 2,296 |  |
| Age (mean (SD)) | 46.43 (16.4) | 47.27 (16.0) | 0.031 |
| Female (%) | 5,567 (54.8) | 1,100 (51.0) | 0.001 |
| **Questionnaire cohort** |  |  |  |
| n | 2,632 | 604 |  |
| Body mass index (median [IQR]) | 25.35 [22.83, 29.03] | 25.47 [23.12, 29.05] | 0.476 |
| Ever smoker (%) | 727 (27.8) | 121 (20.1) | <0.001 |
| Alcohol intake (%) | 2,075 (80.3) | 458 (76.5) | 0.039 |
| Alcohol use (%) | 140 (5.5) | 21 (3.6) | 0.074 |
| Ever used drugs (%) | 123 (4.7) | 12 (2.0) | 0.004 |
| Education level (%) |  |  | 0.842 |
| No formal education | 66 (2.5) | 16 (2.7) |  |
| Primary education | 251 (9.6) | 61 (10.2) |  |
| Secondary education (youth education) | 267 (10.3) | 66 (11.0) |  |
| Vocational training or short-term/medium-term higher education | 1,384 (53.2) | 325 (54.3) |  |
| Long-term higher education | 605 (23.2) | 126 (21.0) |  |
| Unknown | 30 (1.2) | 5 (0.8) |  |

*Alcohol intake: Intake of alcohol within the past 12 months*
*Alcohol use: Reporting >7 units of alcohol a week for females or >14 units of alcohol for males*

Participants were recruited from 13 selected SH areas placed in the municipalities of Copenhagen (n= 5,816), Aarhus (n=3,718), Odense (n=1,731), Hoeje Taastrup (n = 1,308), Helsingoer (n=405) and Slagelse (n= 301), illustrated in supplementary figure 1.

### Seroprevalence

A total of 2,296 (17.3%) participants were seropositive for SARS-CoV-2 antibodies (IgG and/or IgM), of whom 1,594 (69,4%) were positive for IgG antibodies, 1,602 (69,8%) were positive for IgM antibodies and 899 (39.2%) were positive for both IgG and IgM antibodies. This was significantly higher than the seropositivity of 5.8% of the general population (n=22,677, age 41.7 (SD 14.1) years, 48.5% female). Supplementary figure 1 illustrates the seroprevalence in the general population and our study group. The RR range between SH areas varied from 1.1 in Ringerparken in Slagelse to 4.0 in Vollsmose in Odense.

Seropositive participants were older than seronegative participants (47.3 vs 46.4 years, p=0.03) and more likely to be male (p=0.001). Seropositivity stratified on age and gender is shown in figure 2. Both age and gender remained associated with seropositivity in the multiple logistic regression model for gender and place of testing, for an increase in age of 10 years, OR of seropositivity was 1.03 (95% CI 1.00-1.06, p=0.03) and for male gender OR 1.17 (95% CI 1.07-1.29, p=0.001)

**Figure 2:**
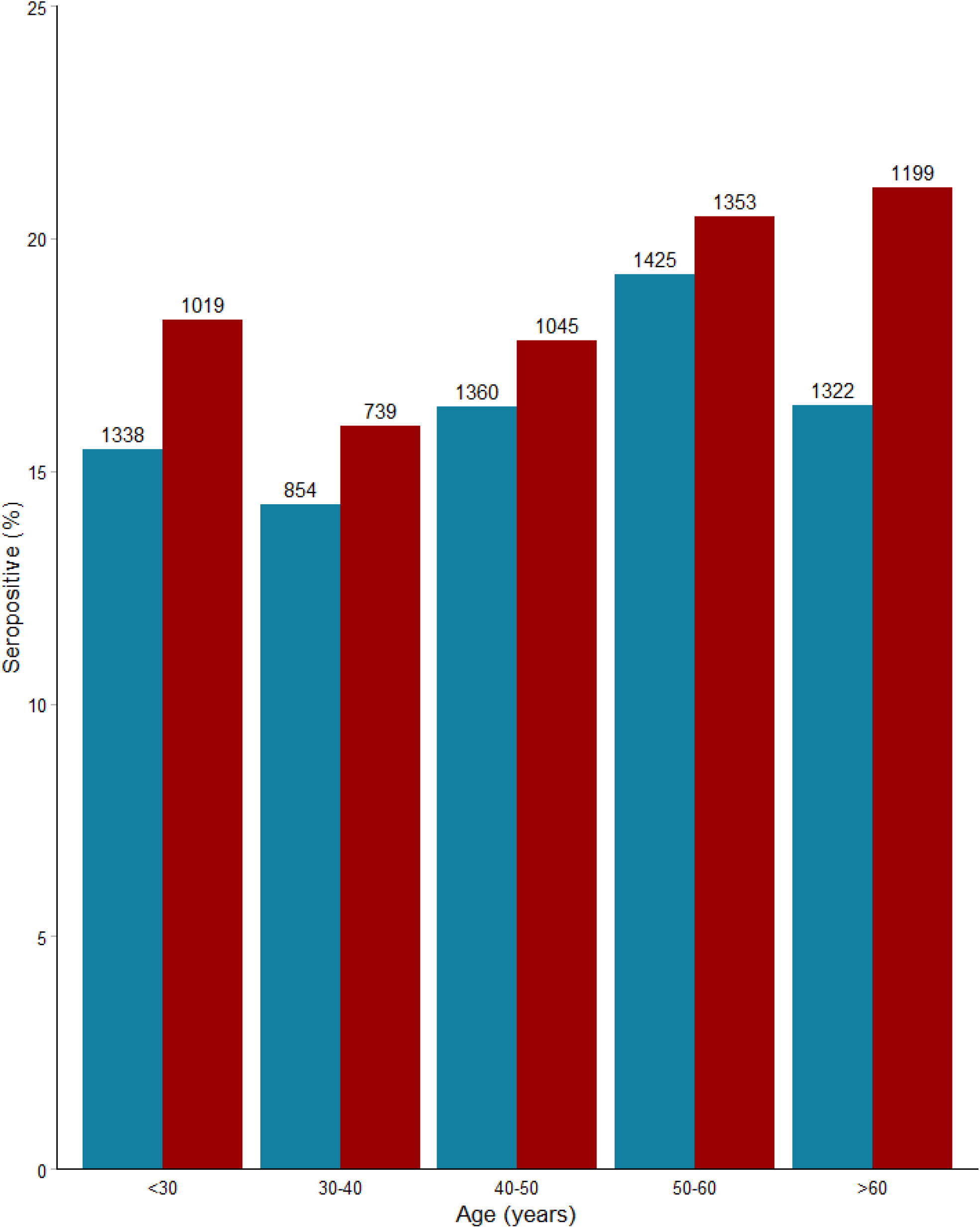
SARS-CoV-2 seroprevalence among 11,654 individuals in SH areas by age and sex *Red: male, blue: female, number of participants in each group*

### Risk factors of infection

Seropositivity stratified by risk factors are shown in table 2. In the analysis adjusted for test location several factors were associated with an increased odds of infection. Reporting a stay of >15 minutes in a room with an infected person (OR 2.7, 95% CI 2.2-3.5, p<0.001), having someone infected in the household (OR 5.0, 95% CI 4.1-6.2, p<0.001), having bodily contact with someone infected (OR 1.2, 95% CI 1.0-1.5, p=0.03) and having a family member with COVID-19 (OR 1.2, 95% CI 1.0-1.5, p=0.03) were all associated with higher odds of infection. Seropositive participants were less likely to smoke (p<0.001), consume alcohol (p=0.04) and engage in drug use (p=0.004) (Table 1). In multivariate logistic regression adjusted for age, gender and place of testing, smoking (OR 0.61, 95%CI 0.48-0.77, p<0.001) and drug use (OR 0.44, CI 0.13-0.26, p=0.01) remained significantly associated with lower odds of seropositivity.

**Table 2:**
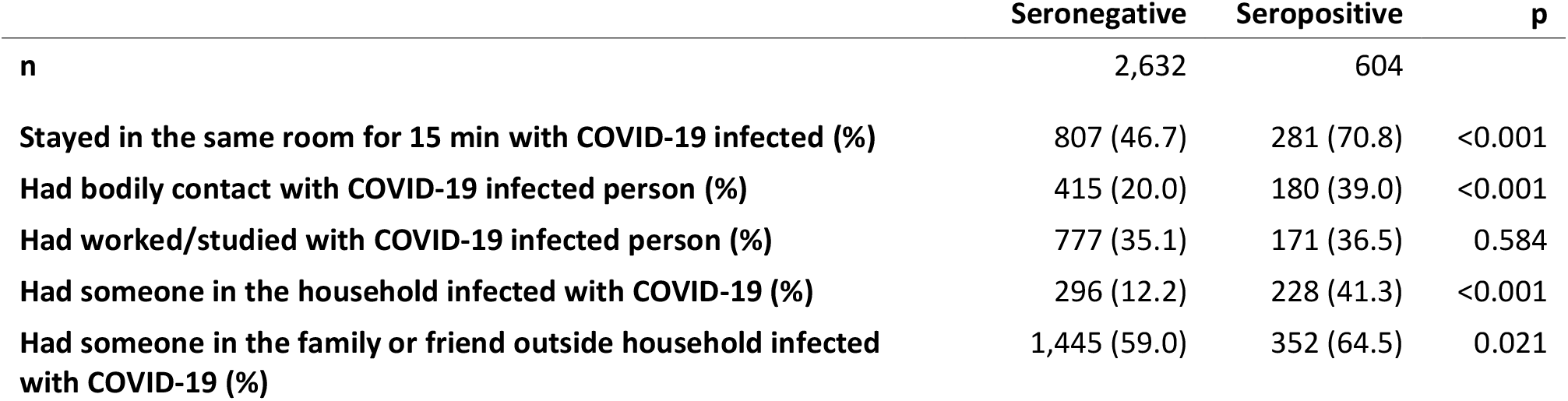
Risk factors stratified by seropositivity of the questionnaire cohort of people in SH areas

Figure 3 shows the risk of seropositivity stratified by the size of the participants household. There was a clear association between the size of households and seropositivity. In adjusted analysis living at least 4 people in a household was associated with a significantly increased risk of seropositivity (OR 1.3, 95% CI 1.1-1.6, p=0.02).Seroprevalence among participants living in a household with only one generation was 16.7%, with two generations 19.2% and with three or more generations 26.1%. Adjusted for age, gender and place of testing, living at least two generations in a household was associated with a significantly increased risk of seropositivity (OR 1.3, 95% CI 1.1-1.6, p=0.003) as compared to living only one generation.

**Figure 3:**
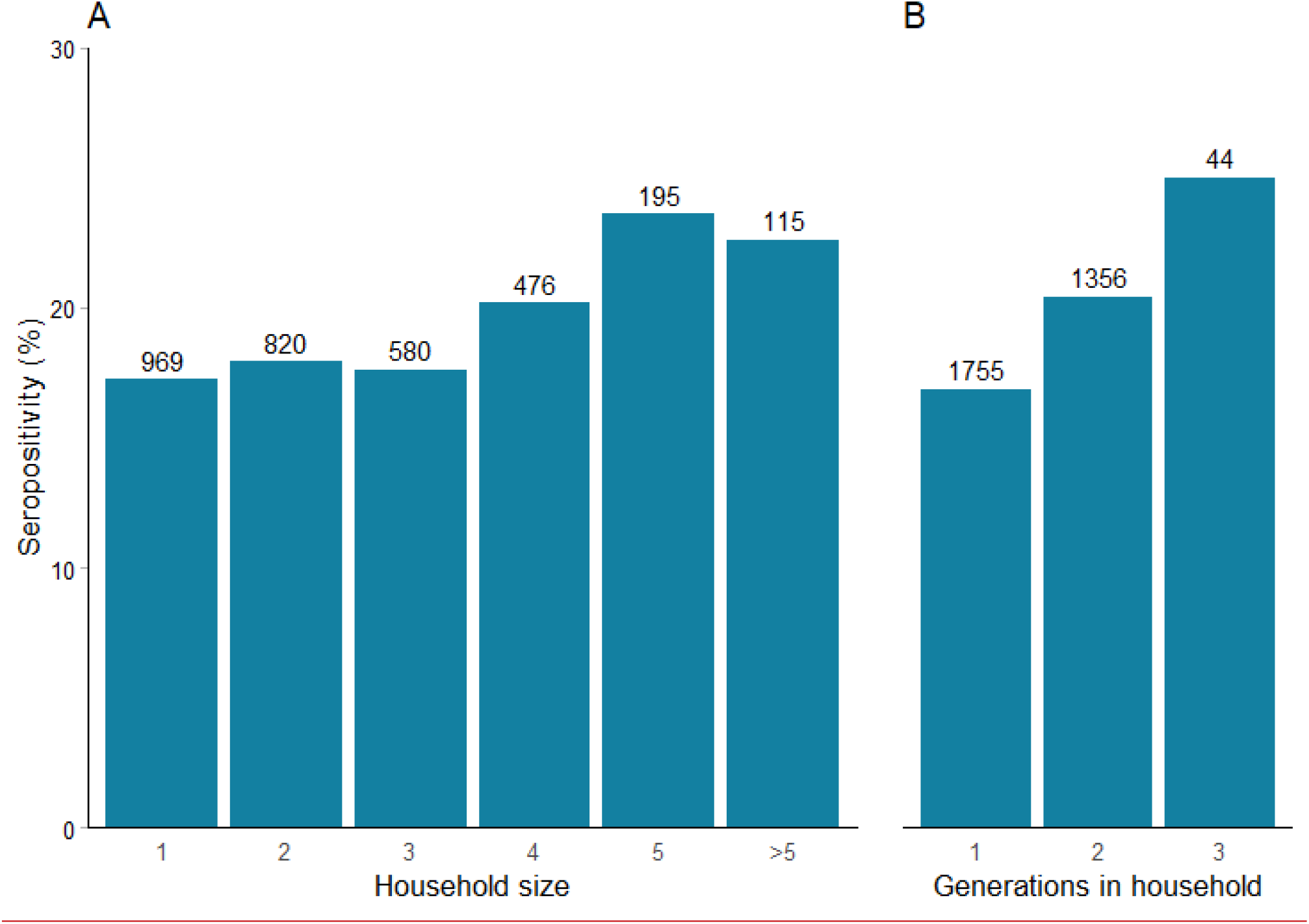
SARS-CoV-2 seroprevalence among 3,155 individuals in SH areas stratified by household size and generations in households *A: Number of household members, B: household size in terms of generations*

Of the 3,236 participants who completed the questionnaire, 2,141 (66.2%) reported working (part time, full time and self-employed participants combined). Figure 4 shows the risk of seropositivity stratified by employment status and occupation. Though nominally increased (19.7% vs 17.0%), in the adjusted model there was no significant increase in seropositivity among participants who reported working (OR 1.2, 95% CI 1.0-1.4, p=0.06).

**Figure 4:**
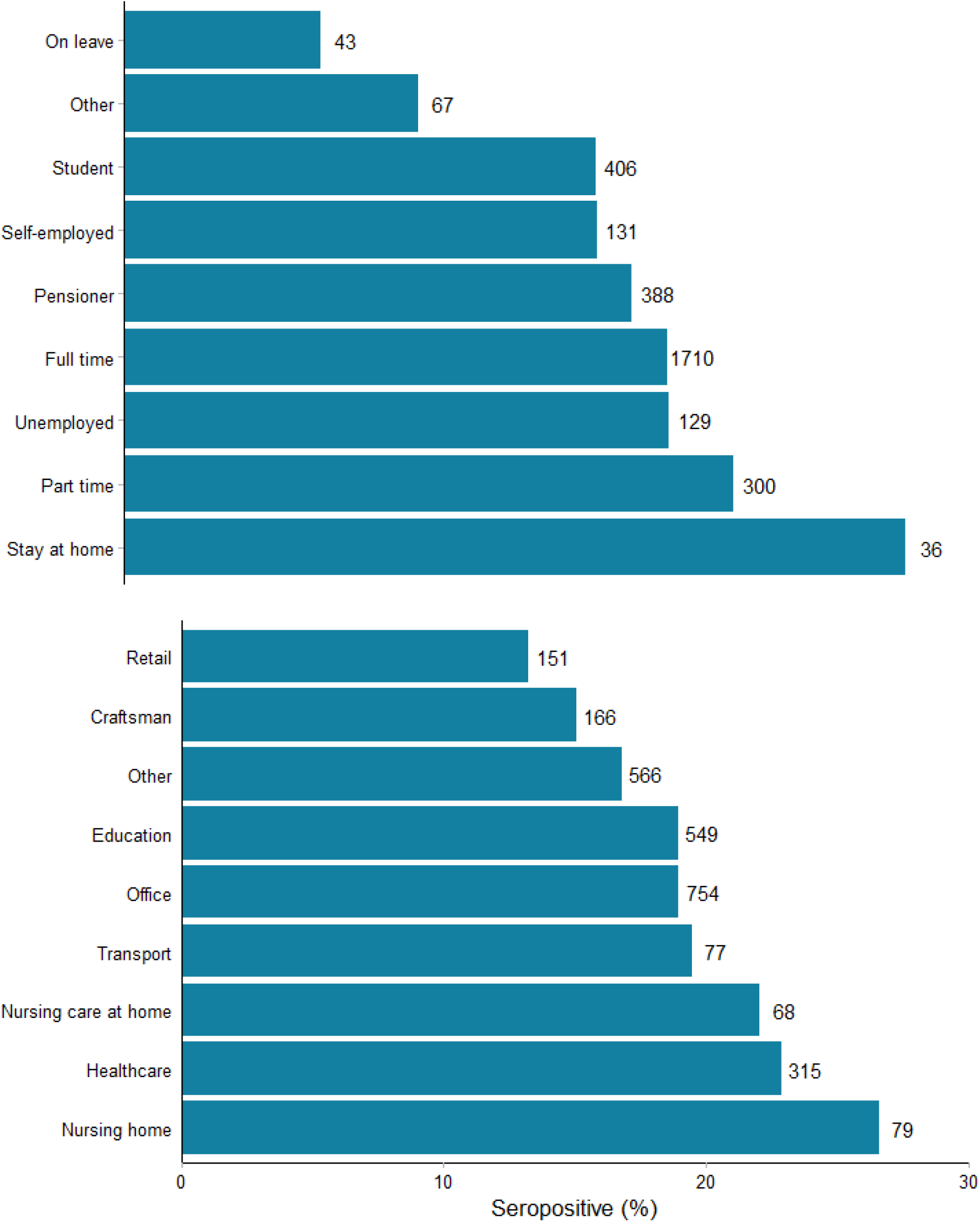
SARS-CoV-2 seroprevalence among 3,210 individuals in SH areas stratified by employment from questionnaire cohort *Upper: relation to job market* *Lower: employment (given participant were employed)* *Categories are as named in questionnaire*.

Seropositivity was numerically higher in participants working at a nursing homes (n=79, 22,9%), participants working in home care nursing (n=68, 22.9%) and participants working in health care (n=315, 22.1%). However, working in health care or at nursing home was not significantly associated to seropositivity compared to the rest of the cohort (OR 1.3, 95% CI 1.0-1.6, p=0.05).

Among the participants who completed the questionnaire, symptoms were reported by 2,355 (72.8%). We found a significant association between reporting of symptoms associated with COVID-19 and seropositivity (RR 1.5, 95% CI 1.3-1.9, p<0.001). Supplementary figure 2 shows the frequency of each symptom stratified for seropositivity. Anosmia (RR 3.2, 95% CI 2.8-3.7, p<0.001) and ageusia (RR 3.3, 95% CI 2.9-3.8, p<0.001) had the strongest correlation to seropositivity. Supplementary figure 3 is a forest plot of risk of seropositivity for each symptom.

### Adherence to the general COVID-19 recommendations from the Danish authorities

The national COVID-19 health recommendations are listed in supplementary figure 4. We found that only 53 (1.6%) participants reported not following any of the national COVID-19 recommendations, listed in the questionnaire (see appendix). Of those 41 were seronegative (77.4%) and 12 seropositive (22.6%), this was not significantly different from other participants (p=0.48). Changes in behavior in response to the COVID-19 epidemic are shown in supplementary figures 5-6. We found that the younger participants were less likely to follow general recommendations. There was no significant association between any behavioral change and seropositivity.

## Discussion

In this large, national, cross-sectional study we determined the seroprevalence of SARS-CoV-2 antibodies in SH areas. The findings from our study indicate that the prevalence of seropositive participants living in SH areas in Denmark was three times higher than in the general population. Males were found to be seropositive more often than females, similar to what has been reported in other studies (12, 13). We found that seroprevalence increased with age, especially among men. Seroprevalence varied between geographical areas, being highest in the Capital region. The prevalence of self-reported symptoms in our study is consistent with previous findings with anosmia and ageusia having the strongest correlation to seropositivity (13, 14). Regarding behavioral factors we saw that seropositive participants were less likely to smoke, drink alcohol or use drugs, this may be related to the fact that older people had higher risk of seropositivity, while alcohol, drugs and smoking is expected to be more widespread among the young. Or related to the fact that people with a higher level of consumption did not participate. We found no significant difference in body-mass index (BMI) between seronegative and seropositive participants, however overall participants were overweight with a median BMI above 25.

In relation to age-related seroprevalence we do not see the same trend as in other Danish studies. The Danish National Seroprevalence Survey of SARS-CoV-2 infection by SSI described seroprevalence estimates were roughly 3 times higher in those aged 12-29 compared to 65 years and above (14) also, a study of 29,295 health-care workers in Denmark found participants younger than 30 years having the highest seroprevalence (13). A study of 1,100 retired blood donors found a lower seroprevalence in the age group >69 years (20), this was also seen in a study of ambulance staff in Sweden and Denmark which described the lowest proportion of seropositive aged 60 years and above (21)

Appropriate quarantine and separation from infected household members can be challenging in large households, and previous studies described how crowded living conditions constitute an increased risk of seropositivity (7, 8, 22, 23). Furthermore, crowded living conditions is considered a key reason why people of low socioeconomic status or from ethnic minority backgrounds in particular have been disproportionately affected by the pandemic (4, 24). Our results show that being exposed to SARS-CoV-2 due to close contact (physical contact and staying in the same room above 15 minutes as infected) or having someone infected in the household increased the risk of seropositivity significantly.

We found that the household composition was of importance, as living at least 4 people in a household or living in a multigenerational household increased the risk of seropositivity among participants. This finding is consistent with a previous preprint study on SARS-CoV-2 transmission within Danish households, which demonstrated an increased transmission risk with age (25).

Joblessness and low levels of education are more common in SH areas than in the general population and a study from Italy reported that being less educated may be a challenge under a pandemic, as the language barrier may affect the adherence to institutional recommendations such as wearing protective masks, avoid contagion and maintaining social distancing (5). In our study only 2,7% seropositive participants answering the questionnaire were without any formal education. Our findings did not indicate clear association between seropositivity and behavioral change overall, and we observed a high rate of behavioral change, especially among the elderly. Only 1.6% of participants indicated not having changed behavior in response to the COVID-19 pandemic, why study participants overall were likely to implement public health measures. Joblessness (stay at home, unemployed or on leave) was not associated with the risk of seropositivity.

Joblessness can potentially be a protective factor against COVID-19 infection as more prone to stay at home and avoiding public transportation or meeting people outside the household (5). Employment in health care or nursing (nursing homes and nursing care at home) was associated with numerically higher seropositivity, however not significantly (p=0,05). This can be due to the fact that these job functions have been less affected by restrictions of working from home and avoiding person-to person contact and thereby pose an increased risk of infection. Also, vaccination of health care workers and workers in nursing homes was introduced December 27^th^, 2020 by the Danish Health Authority and the Ministry of Health as part of the Danish vaccination program, why there is a small likelihood that these participants have been vaccinated and thereby present a positive POCT based on antibodies from the vaccine instead of naturally immunization.

### Ethnicity

A report from the SSI about ethnicity and SARS-CoV-2 infection showed that citizens of non-Western descent were overrepresented by a factor of three in relation to the part they constitute of the Danish population, which may be due to living conditions or employment with higher risk of infection (8). Unfortunately, we could not explore differences by ethnicity, as this information was not available by questionnaire and only a fraction of the participant informed about their social security number with the possibility for obtain information about ethnicity. In future studies like this it will be beneficial to include ethnicity in the questionnaire and include a translator at the test site to prevent potential linguistic barriers in answering of the questionnaires. Selection bias due to language barriers might have affected our results despite the fact that information material and questionnaire were available in seven different languages.

### Strengths and limitations

Our study has several limitations. The study design does not provide information on the point of time when participants became seropositive nor determination of time of infection. It is possible that participants who had been detected with COVID-19 earlier (viral throat-or nasopharyngeal swab or antibody test) or experienced COVID-19 like symptoms did not participate, leading to an underestimation of the seroprevalence and an potential overestimation of participants concerned about prior infection. Seropositivity can be underestimated due to the fact that antibodies in response to SARS-CoV-2 infection can first be detected about one week after symptom onset (22, 26). Seropositivity of Danish blood donors in the same period was used as a proxy for the general population with some limitations to consider. Blood donors were in the age group 17-70 years and our study group are in the age group 15-83 years. Seroprevalence in blood donors is determined on the basis of ELISA and not POCT. In general, blood donors are in good health, why seropositivity in this group could be lower than expected. There is a tendency for health care professionals to be overrepresented as blood donors, and this group is found to have a higher risk of SARS-CoV-2 infection (13), why the seropositivity of blood donors could be higher than expected.Participants did not have to document a residential address in the SH areas, why there may be participants from other areas. There is a small risk of participants from SH areas being vaccinated prior to participation in this study and thereby seropositive based on vaccine response and not natural immunization. Vaccinated blood donors have been deducted from data.

This study had a broad national participation, which give us a representative sample of the Danish population living in SH areas of low socioeconomic status. The high participation rates across the country may reflects a keen interest in knowing the serological status supported by easily accessible testing facilities near the household, and written information in different language. Interobserver variation was limited as the POCT was read by project staff at the test sites. Serological surveys are the best tool to determine the spread of an infectious disease, particularly in the presence of asymptomatic individuals or incomplete ascertainment of those with symptoms. The POCT is a useful serological tool as it is easy to use, provides results in 15 minutes, can be performed by the participants, do not require a venous blood sample nor laboratory equipment and is less costly than ELISA, and thereby a suitable option for large sero-epidemiological studies.

## Conclusion

People living in SH areas in Denmark, have a three times higher seroprevalence of SARS-CoV-2 antibodies compared to the general Danish population. Seroprevalence was significantly higher for males and increased with age. Living in multiple generations or more than four persons in a household was an independent risk factor for being seropositive. Results of this study can be used for future consideration of the need for preventive measures in the populations living in SH areas.

## Data Availability

All personal data obtained in REDCap was kept in accordance with the general data protection regulation and data protection law stated by the Danish Data Protection Agency.

## Contributors

The study was designed and initiated by: KF, RLS and KI

Data analysis was done by: RH and AE

The first draft was written by: KF, AE, RH, HB and KI

All authors have critically revised the manuscript and agree to be accountable for all aspects of the work.

All authors approved the final version of the manuscript.

## Acknowledgements

The authors would like to thank everyone who participated in this study, all staff and local contacts at test sites. We thank Thomas Benfield, Cyril JM Martel and Kåre Mølbak for contributions to this study.

## Figure Legends

**Supplementary figure 1:**
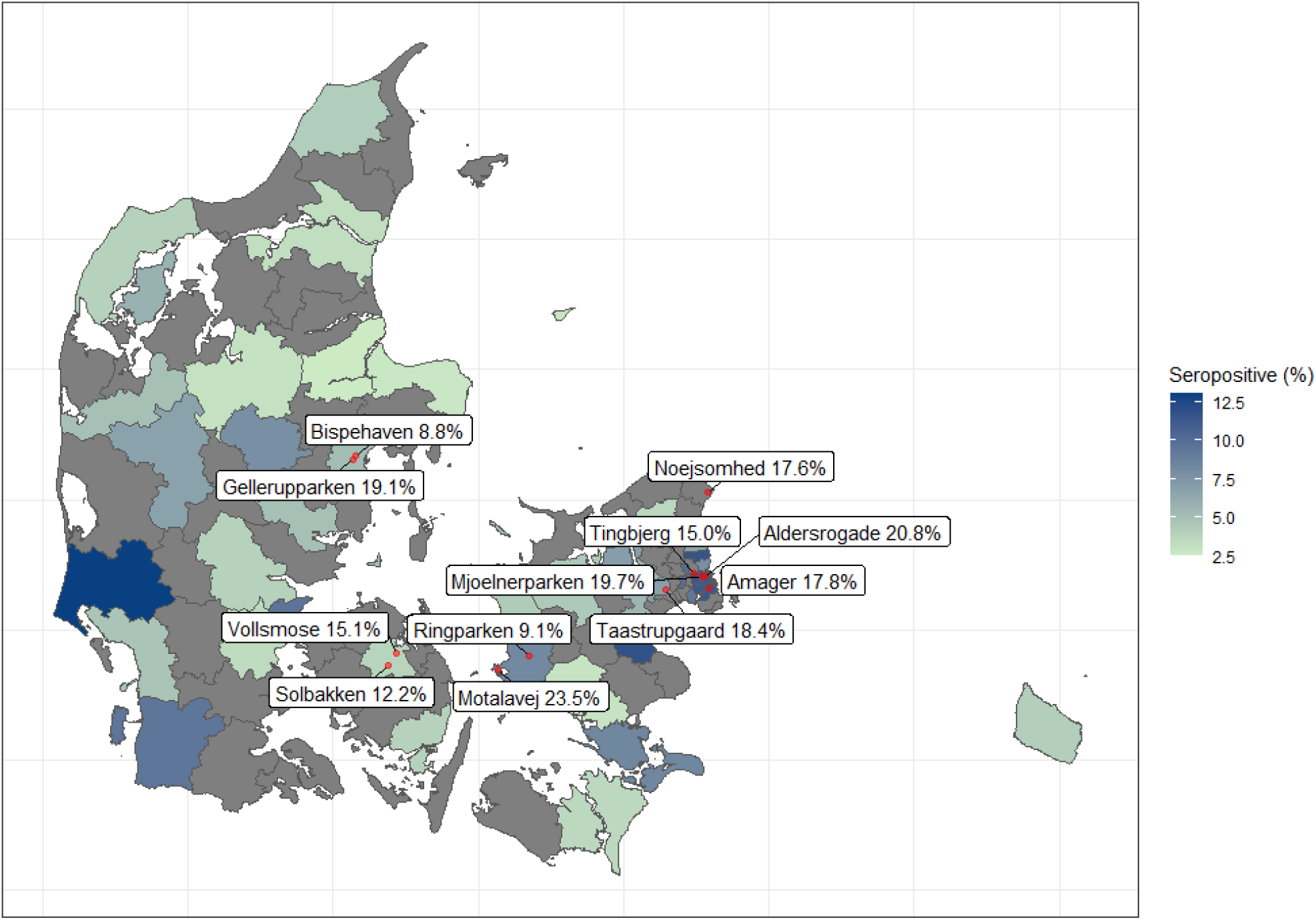
Map of seropositivity *Colors indicating the general population (blood donors)* *Text boxes indicating the study cohort.* *Gellerupparken: Including SH area Gellerupparken and Bazar Vest* *Noejsomhed: Including SH area Noejsomhed and SH area Vapnagaard*

**Supplementary figure 2:**
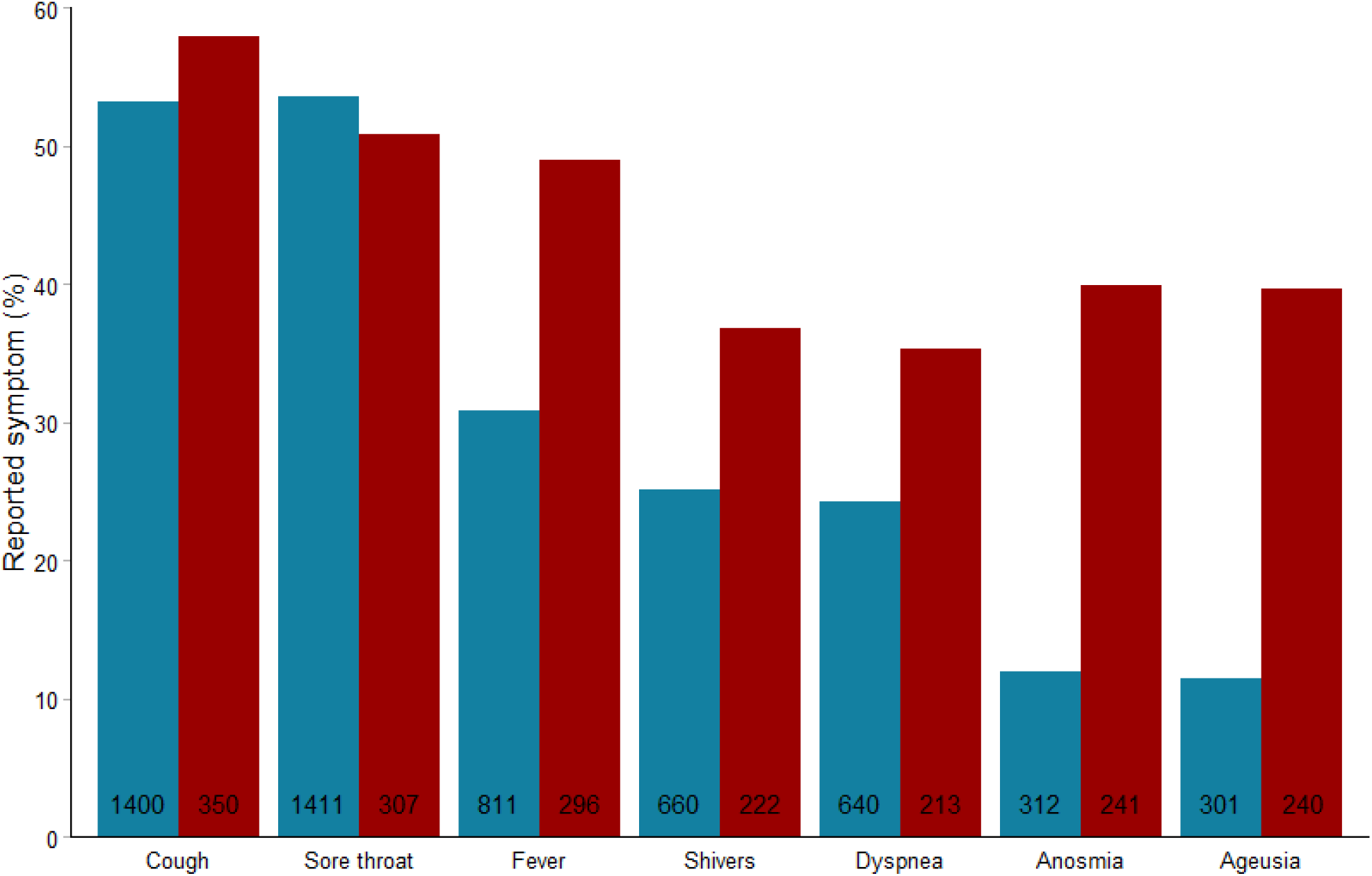
Frequency of symptoms among 3,236 individuals in SH areas stratified by seropositivity *Red: Seropositive, Blue: Seronegative*

**Supplementary figure 3:**
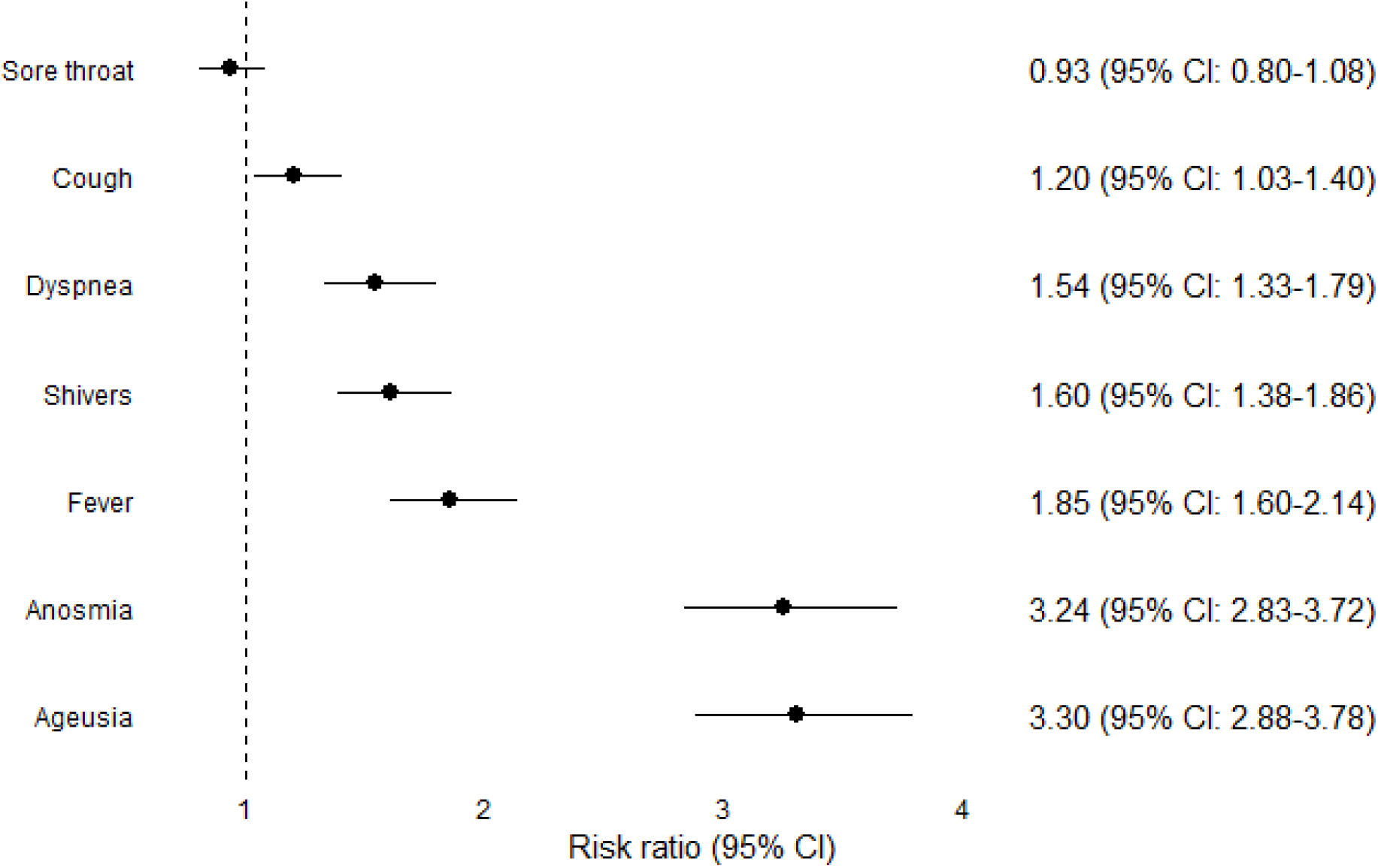
Forest plot of risk ratios (RR) for each symptom reported by questionnaire cohort

**Supplementary figure 4:**
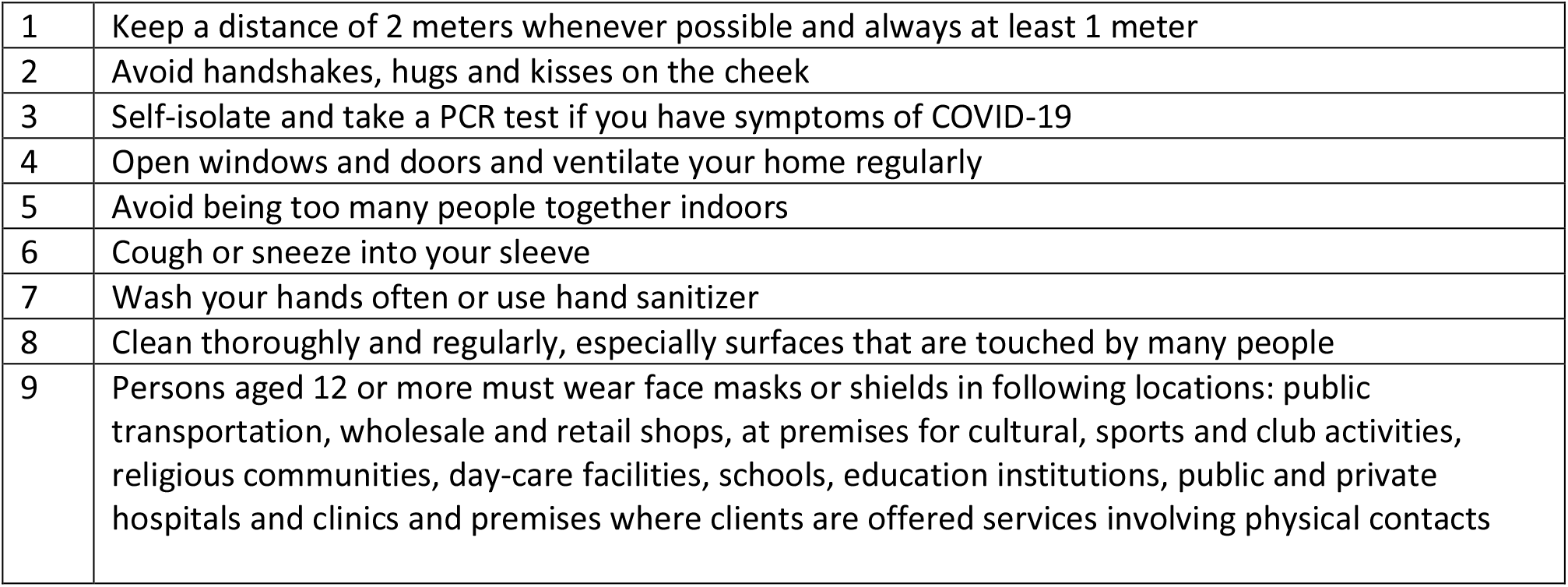
General recommendations from the Danish Health authorities

**Supplementary figure 5:**
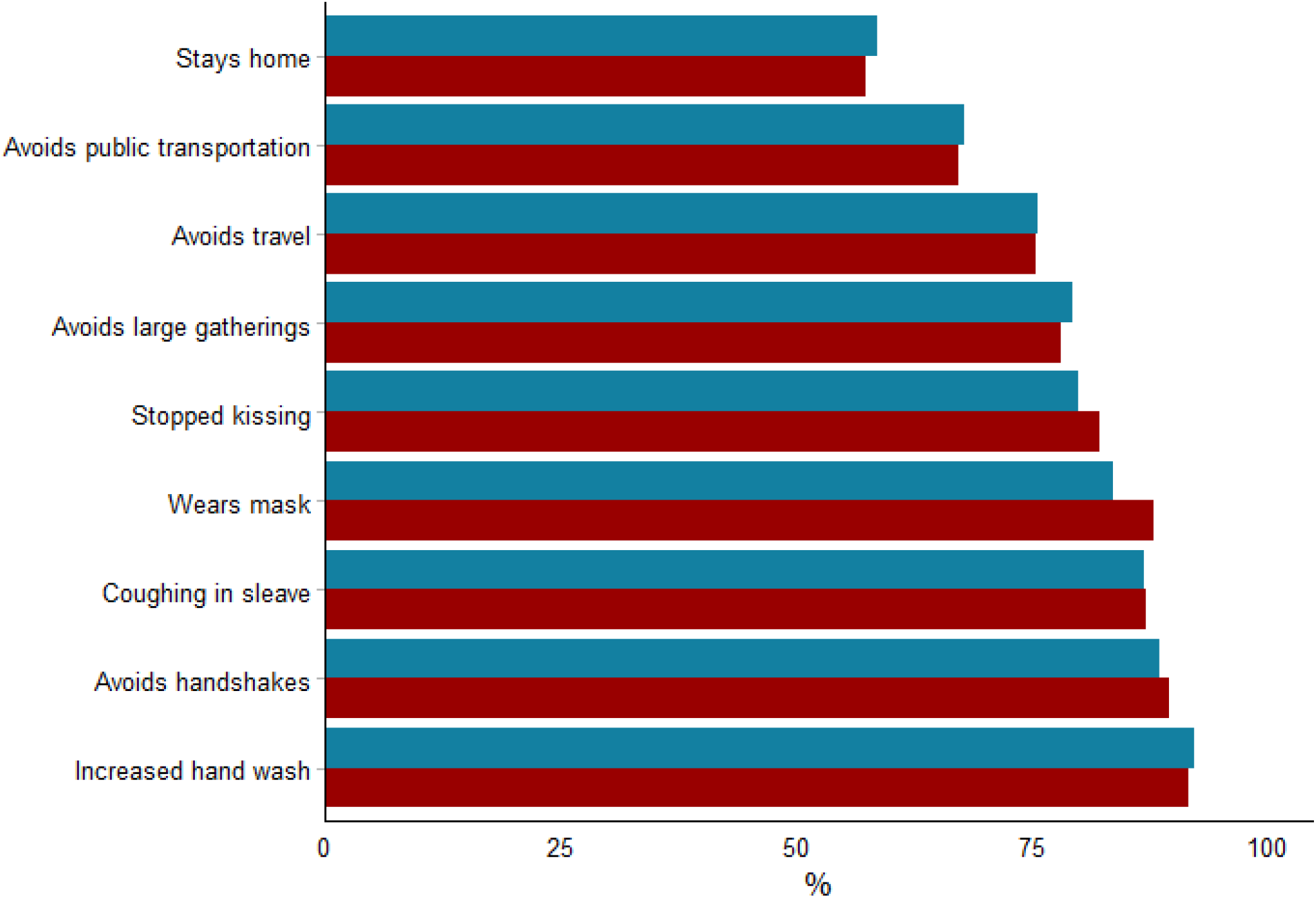
Change of behavior among 3,236 individuals in Danish SH areas during the pandemic stratified by seropositivity *Red: seropositive, Blue: seronegative*

**Supplementary figure 6:**
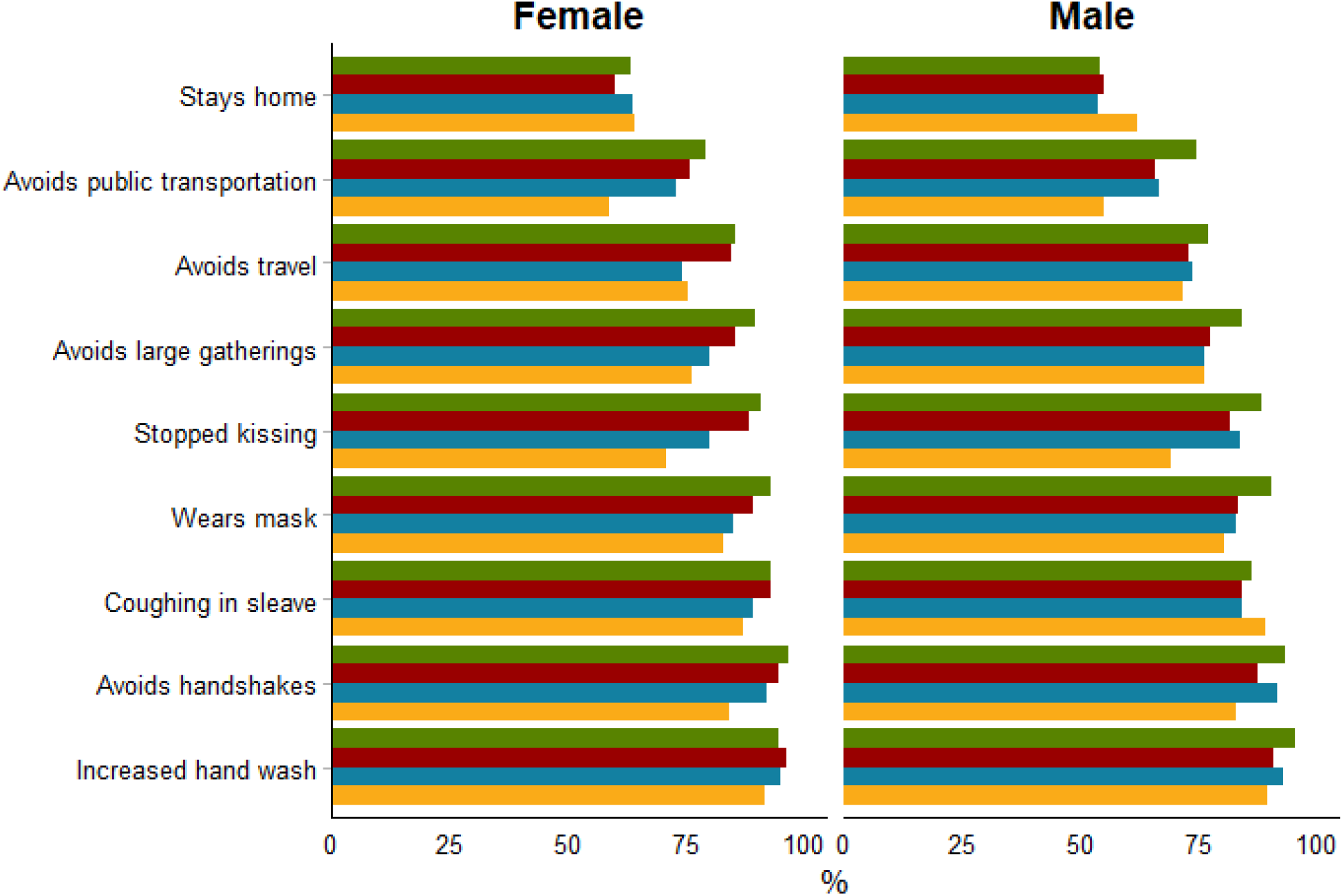
Change of behavior among 2,871 individuals in Danish SH areas during the pandemic stratified by sex and age in quartiles *(Green – age >56 years, red – age 45-56 years, blue – age 31-45 years, yellow -<31 years)*

## Appendix

### List of test sites

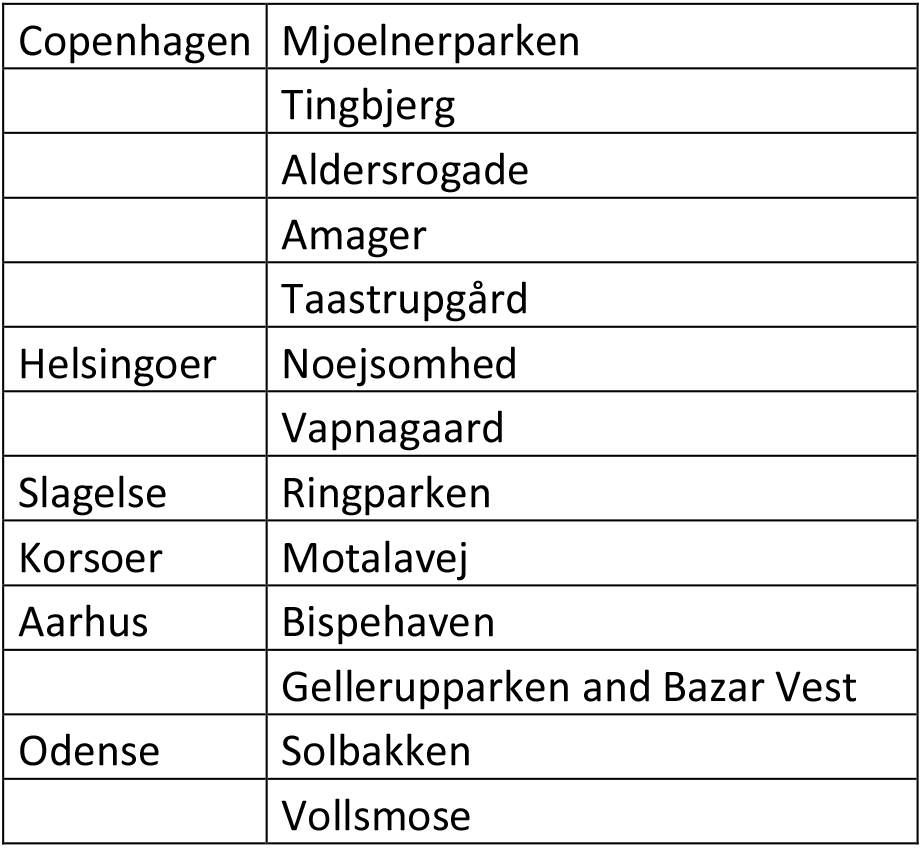

### The questionnaire

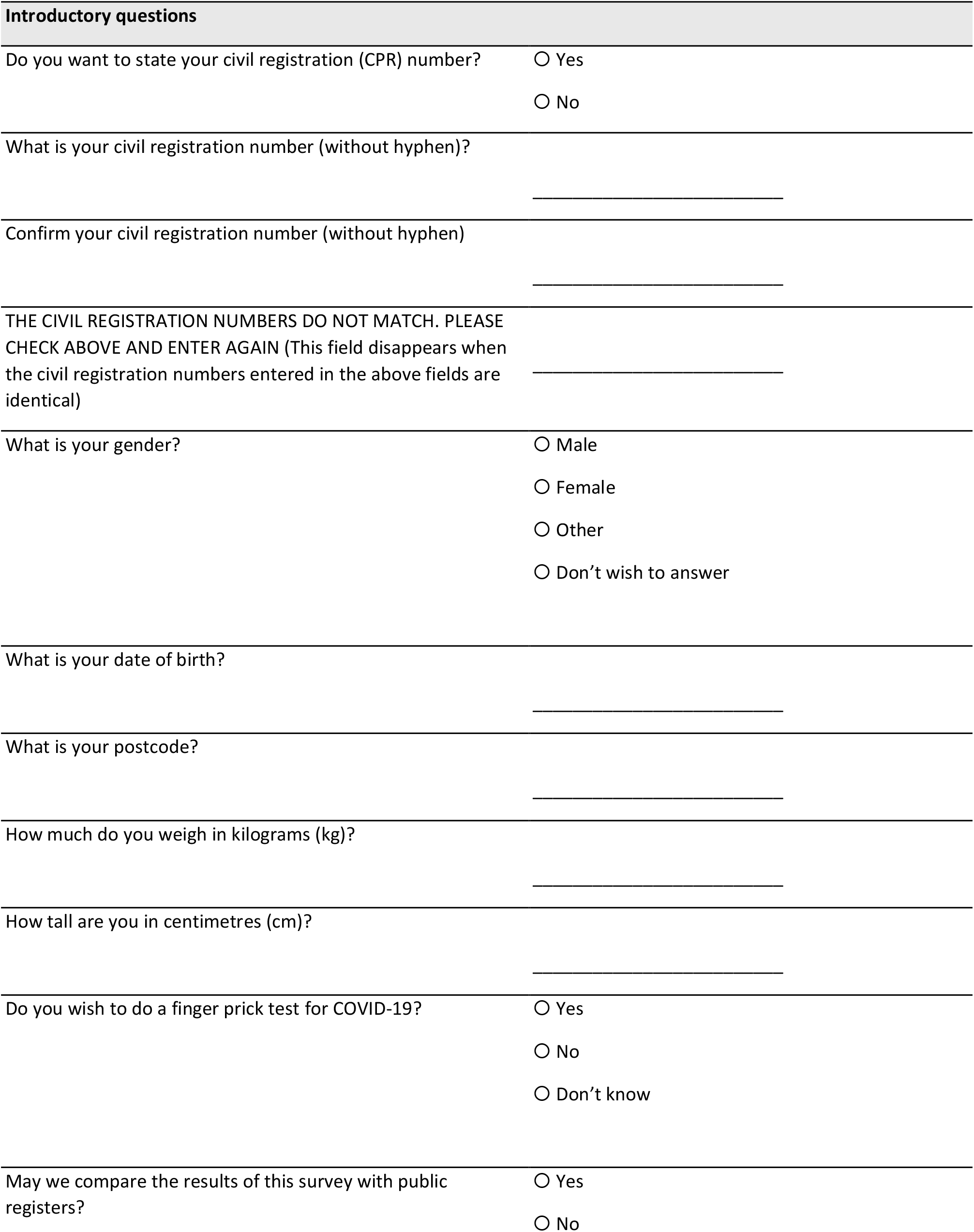
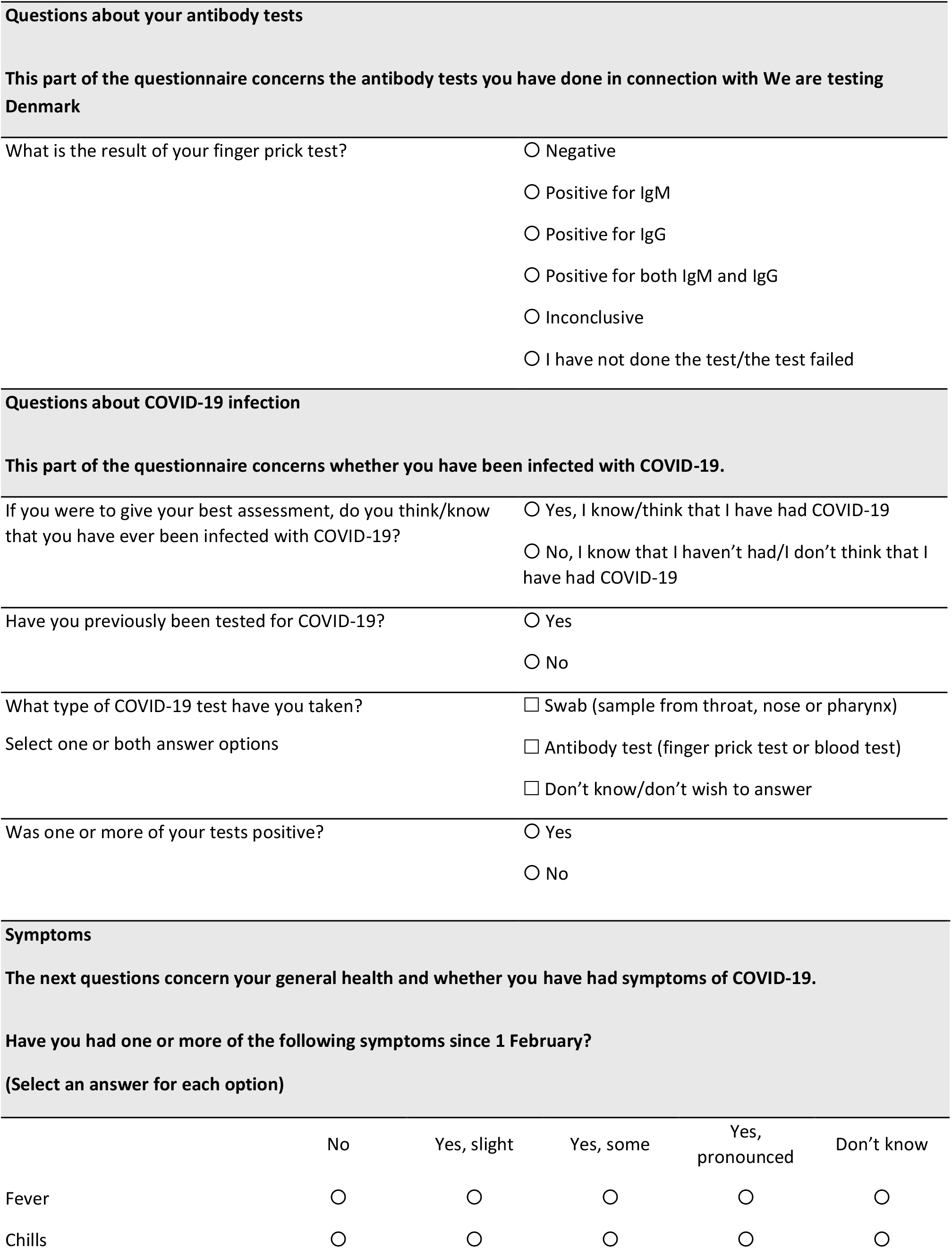
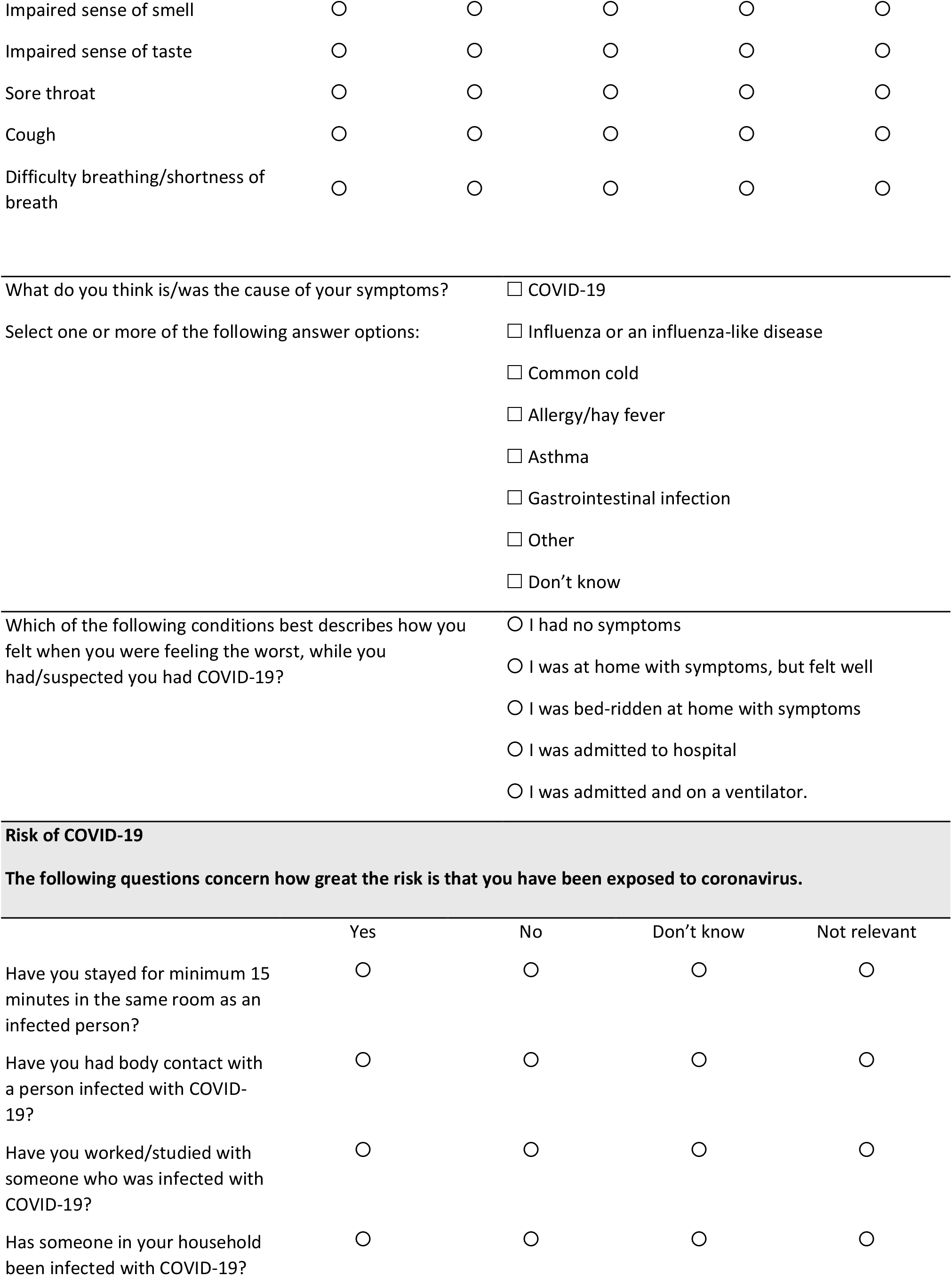
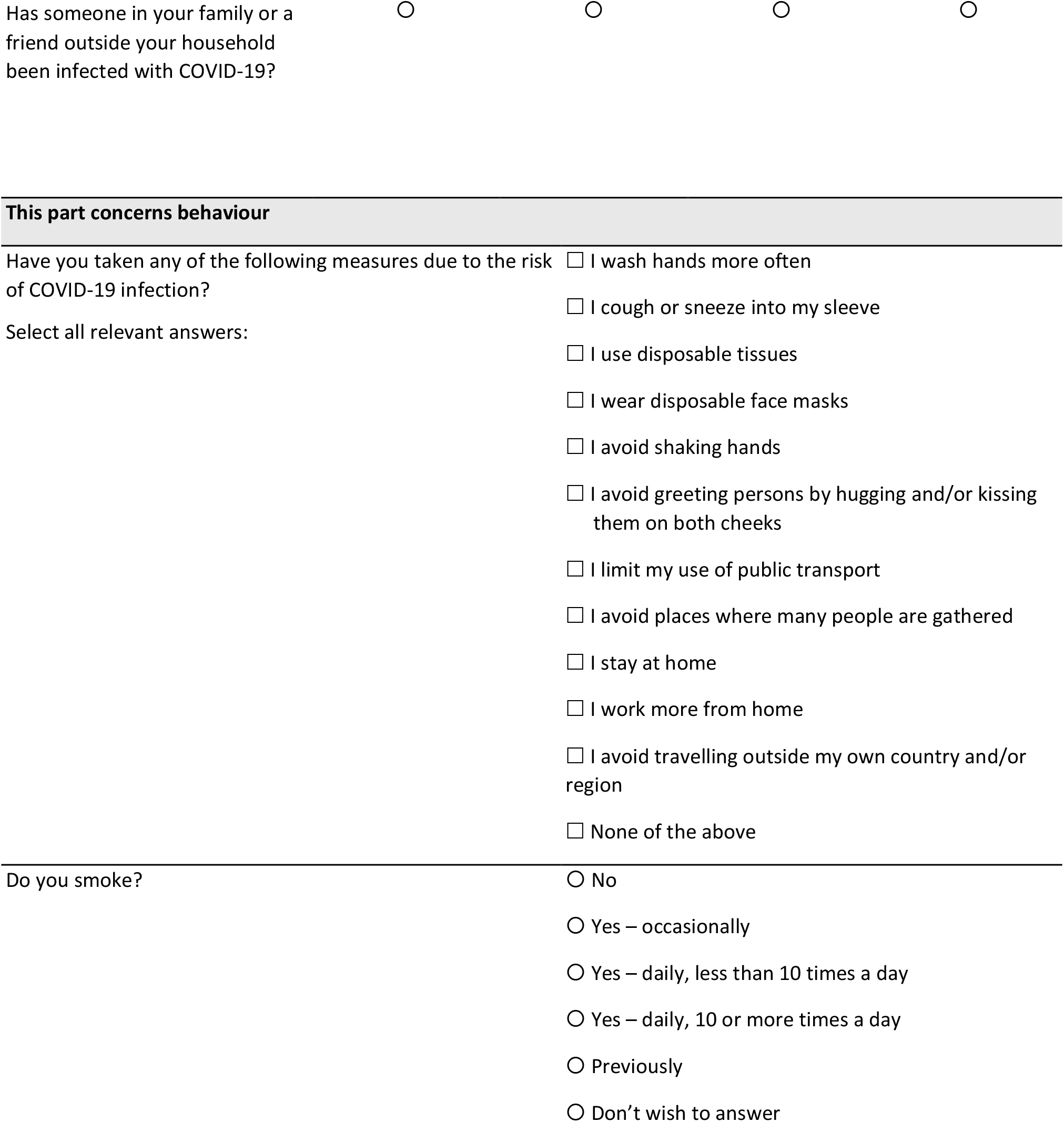
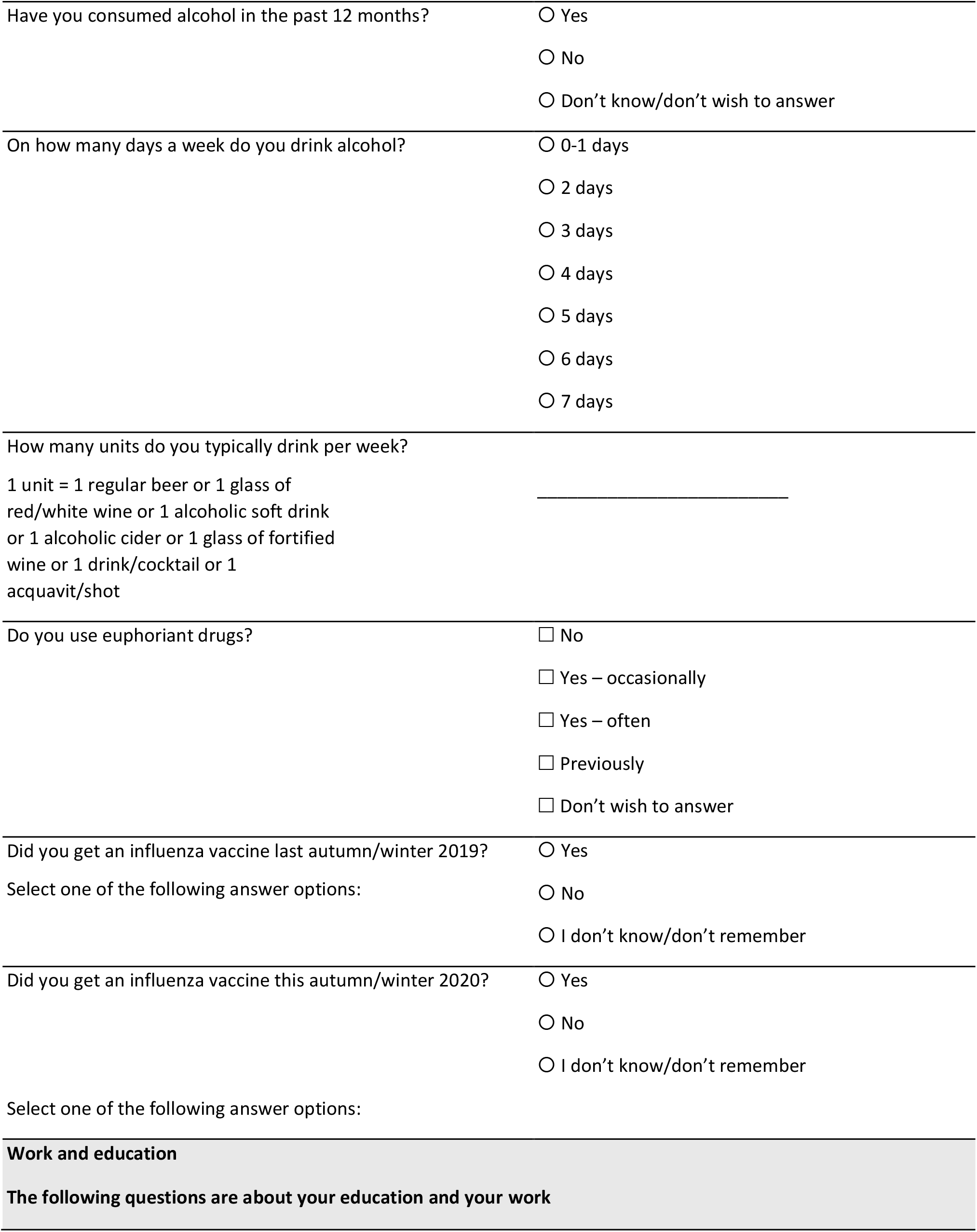
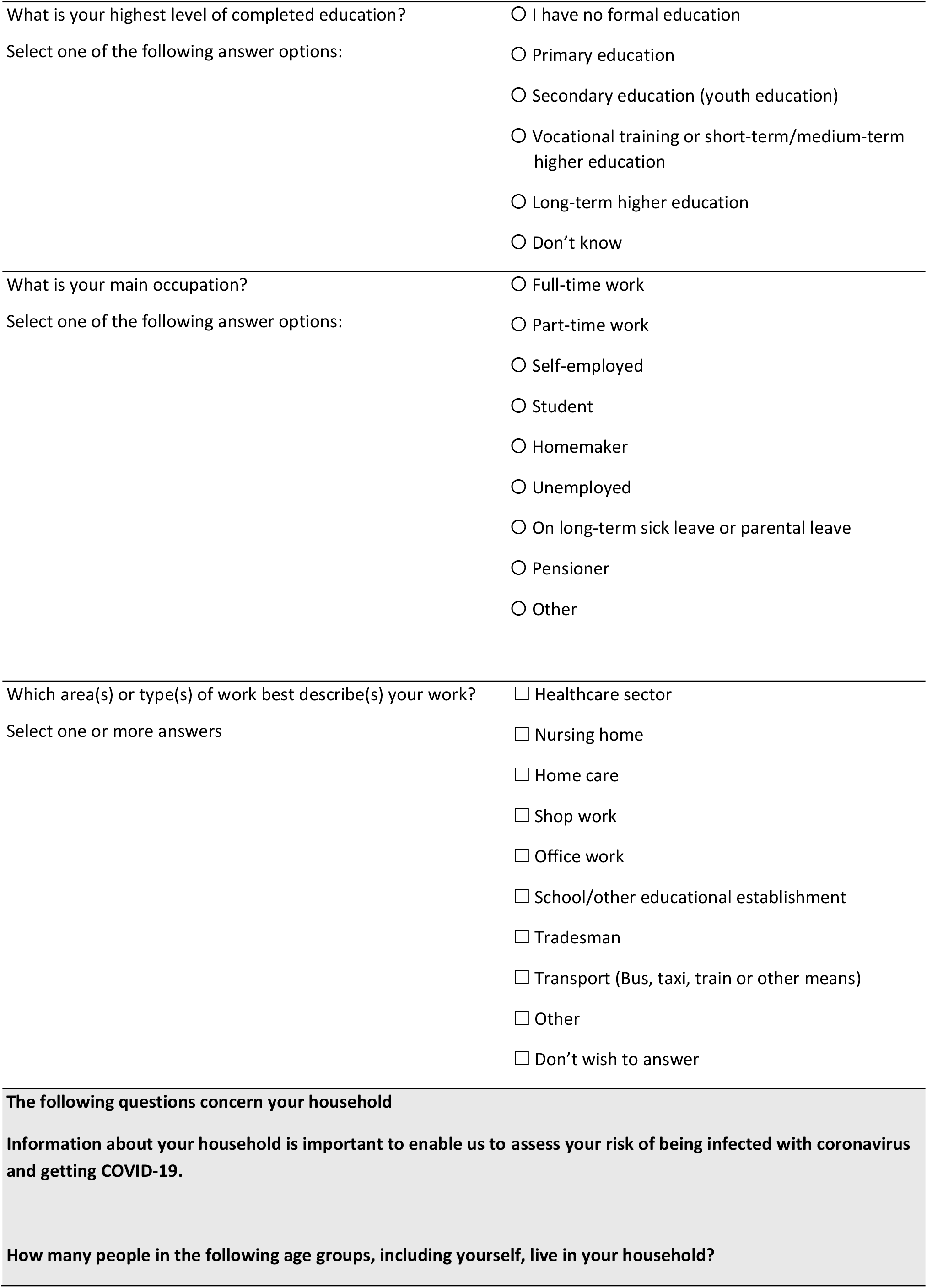
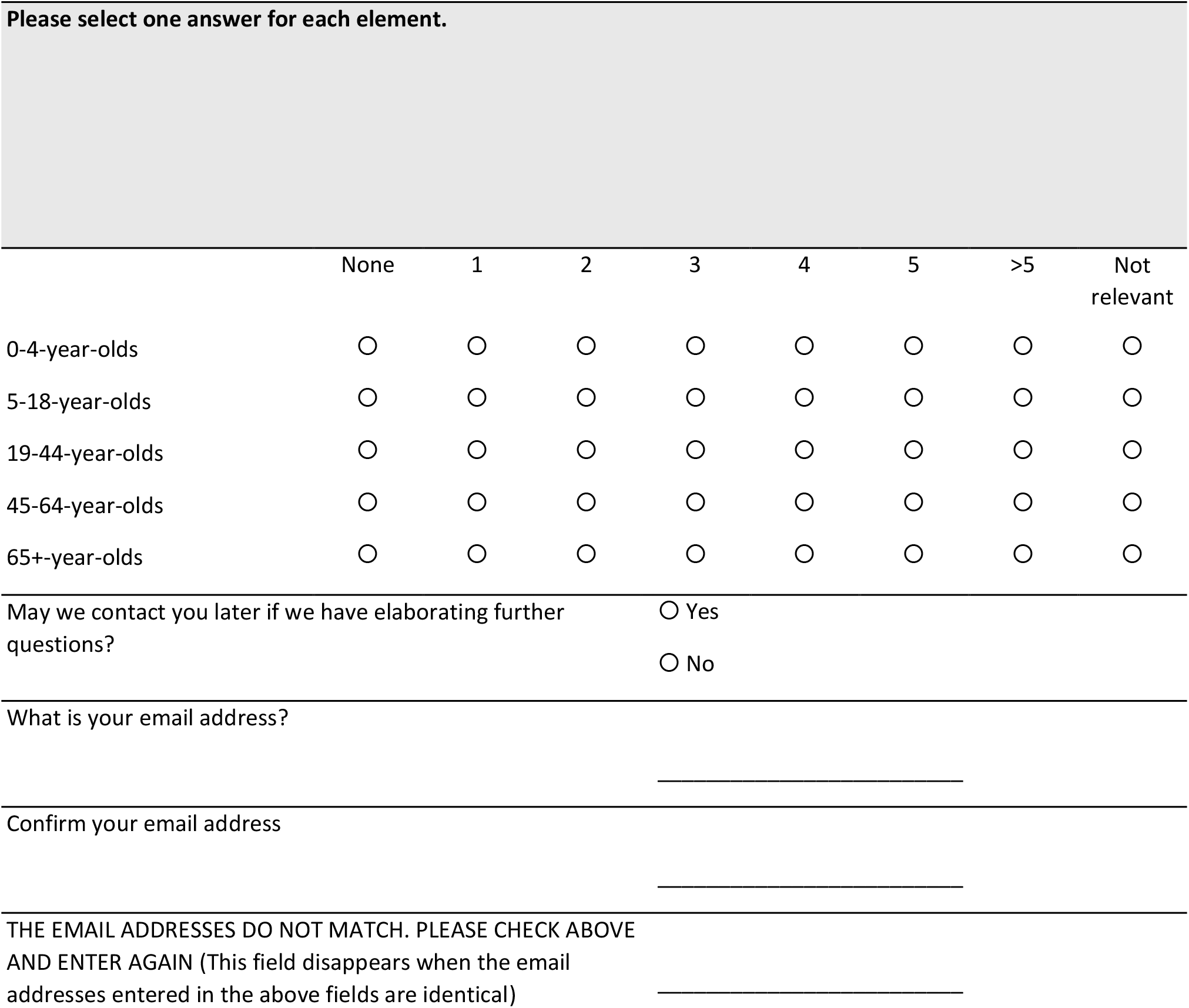

